# Cohort profile: Actionable Register of Geneva Out- and inpatients with SARS-CoV-2 (ARGOS)

**DOI:** 10.1101/2021.05.24.21256813

**Authors:** Camille Genecand, Denis Mongin, Flora Koegler, Dan Lebowitz, Simon Regard, Mayssam Nehme, Olivia Braillard, Marwène Grira, Dominique Joubert, Pierre Chopard, Elisabeth Delaporte, Jerome Stirnemann, Idris Guessous, Aglaé Tardin, Delphine S. Courvoisier

## Abstract

**Purpose:** The Actionable Register of Geneva Out- and inpatients with SARS-CoV-2 (ARGOS) is an ongoing prospective cohort created by the Geneva Directorate of Health (GDH). It consists of an operational database compiling all SARS-CoV-2 test results conducted in the Geneva area since late February 2020. This article aims at presenting this comprehensive cohort, in light of some of the varying public health measures in Geneva, Switzerland, since March 2020.

**Participants:** As of June 1st, 2021, the database included 356’868 patients, among which 65’475 had at least one positive test result for SARS-CoV-2. Among all positive patients, 37.6% were contacted only once, 10.6 % had one follow-up call, 8.5% had two, and 27.7% had 3 or more follow-up calls. Participation rate among positive patients is 94%. Data collection is ongoing.

**Findings to date:** ARGOS data illustrates the magnitude of COVID-19 pandemic in Geneva, Switzerland, and details a variety of population factors and outcomes. The content of the cohort includes demographic data, comorbidities and risk factors for poor clinical outcome, self-reported COVID-19 symptoms, environmental and socio-economic factors, prospective and retrospective contact tracing data, travel quarantine data, and deaths. The registry has already been used in several publications focusing on symptoms and long COVID, infection fatality rate, and re-infection.

**Future plans:** The data of this large real-world registry provides a valuable resource for various types of research, such as clinical research, epidemiological research or policy assessment as it illustrates the impact of public health policies and overall disease burden of COVID-19.

**STRENGTHS AND LIMITATIONS OF THIS STUDY:**

- ARGOS’ main strength consists of its large number of cases, representative of all diagnosed cases on a regional level with the primary aim of assessing all cases.
- ARGOS involves every individual who performed a SARS-CoV-2 test (PCR or antigenic) and is not limited to hospitalized patients, thus providing a valuable resource to assess the overall disease burden of COVID-19 in a geographically defined population.
- To mitigate confounding effects and improve data analysis and interpretation, we present the data according to four policy periods.
- This cohort is multicentric as it includes all tests performed in Geneva’s hospitals (both public and private), private practices and medical centers.
- Due to operational needs, symptoms and comorbidities are self-reported, which may lead to measurement error or misclassification.

## Introduction

In December 2019, an increasing number of cases of pneumonia caused by a novel coronavirus, SARS-CoV-2^1^, was observed in Wuhan, China. On March 11, 2020, the World Health Organization (WHO) declared the coronavirus disease 2019 (COVID-19) outbreak a global pandemic(1,2). As of June 1st, 2021, the virus spread to 207 countries, infected close to 171 million people and caused 3.68 million deaths(3,4). In Switzerland, the cumulative incidence of laboratory confirmed COVID-19^2^ cases during the first wave was in the top five countries in Europe, with about 400 confirmed cases per 100’000 population at the end of July 2020(4,5). In the Geneva area, the first COVID-19^2^ patient was diagnosed on February 26, 2020(6). Possibly due to the city’s geographical proximity to Northern Italy(7), the epidemic curve showed a steep upward trend. The first wave of the epidemic peaked in Geneva on April 2nd with 233 cases in 24 hours in an area with a population of 500’000. Geneva’s cumulative incidence of confirmed cases is almost 3 times that of Switzerland(5), with more than 1’000 cases per 100’000 population(6), while the seroprevalence was estimated to be close to 10 times that of the confirmed cases as 9.7% of the population had antibodies three weeks after the height of the epidemic(8,9).

A database was created in early March in order to contact new cases and keep track of their follow-up. The Actionable Register of Geneva Out- and inpatients with SARS-CoV- 2^1^ (ARGOS) includes all SARS-CoV-2^1^ test results conducted in the Geneva area since late February 2020, as well as those from Geneva residents being tested in other Swiss cantons. After more than a year of pandemic and guided by operational needs, ARGOS has been enriched by various data, including contact tracing information. The primary aim of this article is to present this comprehensive cohort, its characteristics and the content of the data collected. The secondary aim is to interpret the data according to the public health measures implemented over time since the cohort profile was influenced by the varying policies enacted by the Swiss government and the Geneva State.

## Cohort description

### The ARGOS database

ARGOS is an ongoing prospective cohort created by the Geneva Directorate of Health (GDH) and consists of an operational database compiling all SARS-CoV-2 test results conducted in the Geneva canton. Data are collected and managed using the REDCap electronic data capture tools(10,11) allowing the GDH to contact positive cases in order to promote public health measures and coordinate medical follow-up. It is set up as a collaborative tool between different institutions and medical entities, including the GDH, Geneva University Hospitals (HUG), and Geneva’s main private medical centers. The latter have restricted access to data regarding their own patients only. The GDH and HUG are the only users to implement follow-up data in the electronic register. The data is hosted on HUG’s secure servers. The register is administered by a committee of co-Principal Investigators belonging to the GDH and HUG, with the agreement of the cantonal ethic committee (CCER protocol 2020-01273). Participants in the database had the opportunity to refuse to participate in the registry, and those who did are excluded from the analyses presented here and any data sharing. The participation rate for positive patients is 93.9% (calculated as the ratio between the number of patients who gave their consent for the reuse of their data and the total number of patients). As recommended by the World Health Organization, deidentified ARGOS data are made available upon reasonable request, including a research protocol, using the <u>form</u> https://edc.hcuge.ch/surveys/?s=TLT9EHE93C.

### Data collection

All Geneva laboratories performing SARS-CoV-2 testing are required to send the results to the GDH. Swabs are collected from the upper respiratory tract in medical centers, private practice or during home visits by trained healthcare professionals(12). Between January 24, 2020 and June 1st, 2021, 655’464 tests for SARS-CoV-2 recorded in the ARGOS database, 584’512 were performed by real-time reverse transcriptase– polymerase chain reaction assays and 70’952 by rapid antigen tests. The majority were performed in the Geneva area and a small number consisted of tests conducted on Geneva residents in other Swiss Cantons, and declared to the GDH by the Federal Office of Public Health (FOPH). Importantly, patients reporting COVID-19 symptoms between March 13 and March 29, 2020, did not get tested due to shortage of testing materials, unless they were healthcare workers, considered at-risk or hospitalized. However, symptomatic patients who visited the HUG COVID-19 testing center without fulfilling testing criteria were entered in the database as “suspected cases”. Some of these patients later received a test as policy evolved on March 30, 2020. For each positive or suspect case, a series of surveys is filled using REDCap platform. Depending on the needs, follow-up calls are performed either by a professional nurse, a medical student or a contact tracer with supervision from a medical doctor. Findings are documented in the database. 669 patients from the cohort were also called back at 6 week and 7 months to monitor the persistence of symptoms, of which 510 and 410 answered respectively. All SARS-CoV-2 positive patients in Geneva who require hospitalization at HUG received follow-up calls by the HUG team at the time of discharge from the hospital. COVID-19 positive patients identified as nursing home residents or who are hospitalized at the time of diagnosis are not systematically called since they already receive medical attention and isolation measures are enforced by the medical staff. During the second wave, which started in late September 2020, the incidence of SARS-CoV-2 positive patients became so high that the GDH team could not contact everyone in time. A semi-automatic process was put in place. Positive patients and their declared contacts received an invitation to an online survey where they filled basic information. Only then and when the workload allowed it, they received a phone call from the GDH team to complete the data already provided. At the peak of the second wave, not all SARS-CoV-2 positive patients could be contacted. Follow-up calls as well as calls to close contacts were also temporarily abandoned. Finally, the Geneva Cantonal Population Office are required to declare COVID-19 related deaths, which are also recorded in ARGOS. Patients or the public were not involved in research.

### What is being measured?

An overview of collected data is provided in Table 1. The surveys were created by the GDH and HUG medical task forces. Within the first 48h of testing, patients with a positive test result for COVID-19 receive a call by a professional nurse or a trained contact tracer with support from a medical doctor if needed. During this call, demographic data are collected (13), as well as symptoms (14–17), clinical and environmental risk factors, and clinical red flags. A special attention is paid to psychosocial and cultural factors, and resources are provided when needed. The clinical evaluation is used to identify patients who need immediate emergency care, or to address them for follow-up care by their general practitioner, by one of Geneva’s medical centers, or by the GDH-HUG team via telemedicine, which is recorded in the database as well. Patients’ declared symptoms are recorded in subsequent surveys. Patients’ self-reported compliance to isolation measures are also recorded. As of April 27, 2020, close contacts of index cases are individually contacted and basic information is recorded. Demographics, the type of contact they had with the index case, vaccine information, the presence of COVID-19 symptoms and their compliance to quarantine measures are also recorded at first call and during follow-up calls. Since July 6, 2020, the FOPH has established an evolving red list of countries where incidence rate is considered high or with variant of concern. Travelers who stayed in one of these countries have to quarantine for 10 days at their arrival in Switzerland. People staying in Geneva must self-declare upon arrival and fill an online survey containing basic information which data is also part of ARGOS. Depending of the work load, travelers are called by contact tracers during their quarantine period. Self-reported compliance to quarantine measures and the presence of symptoms are recorded during these calls.

**Table 1:**
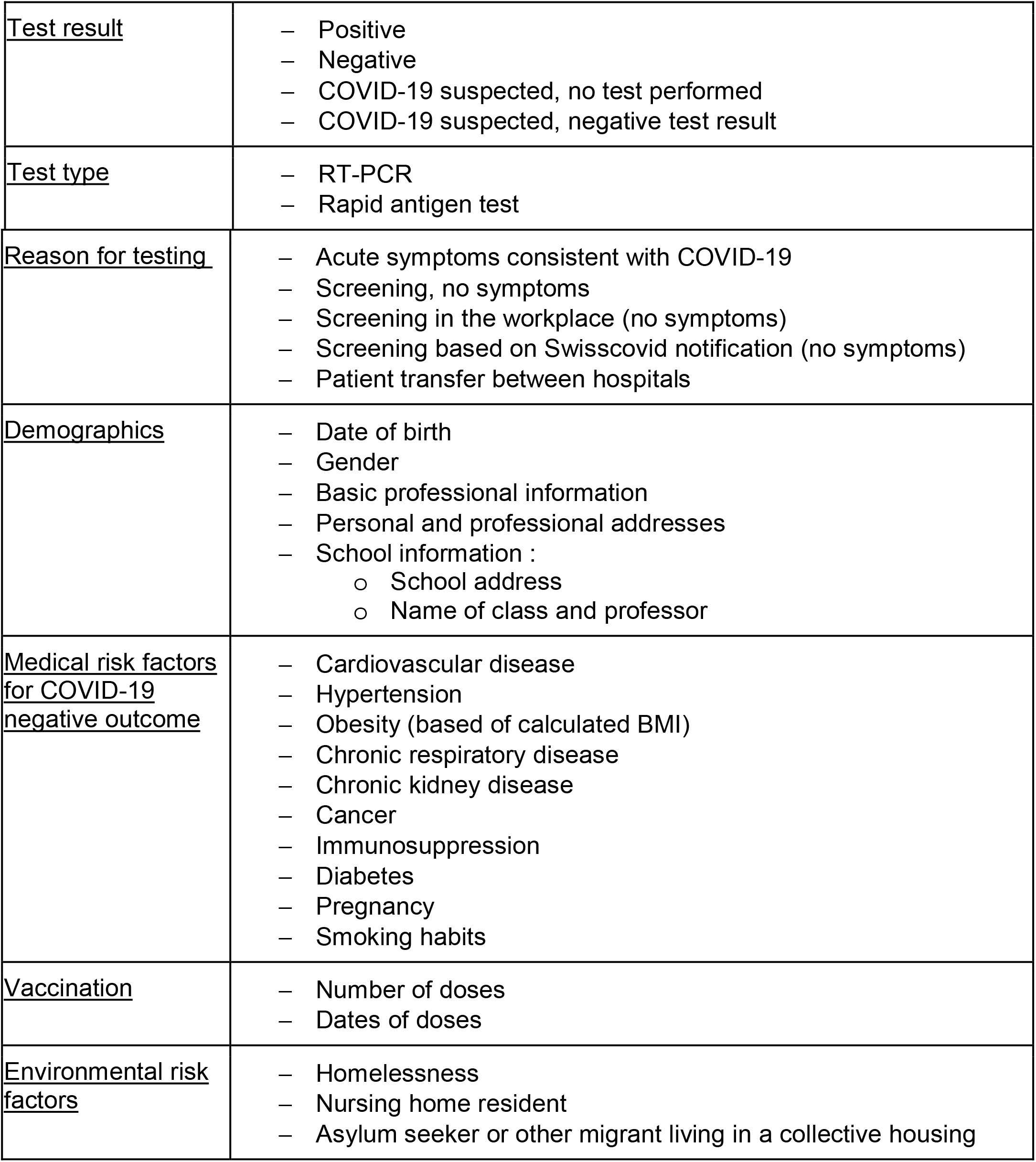
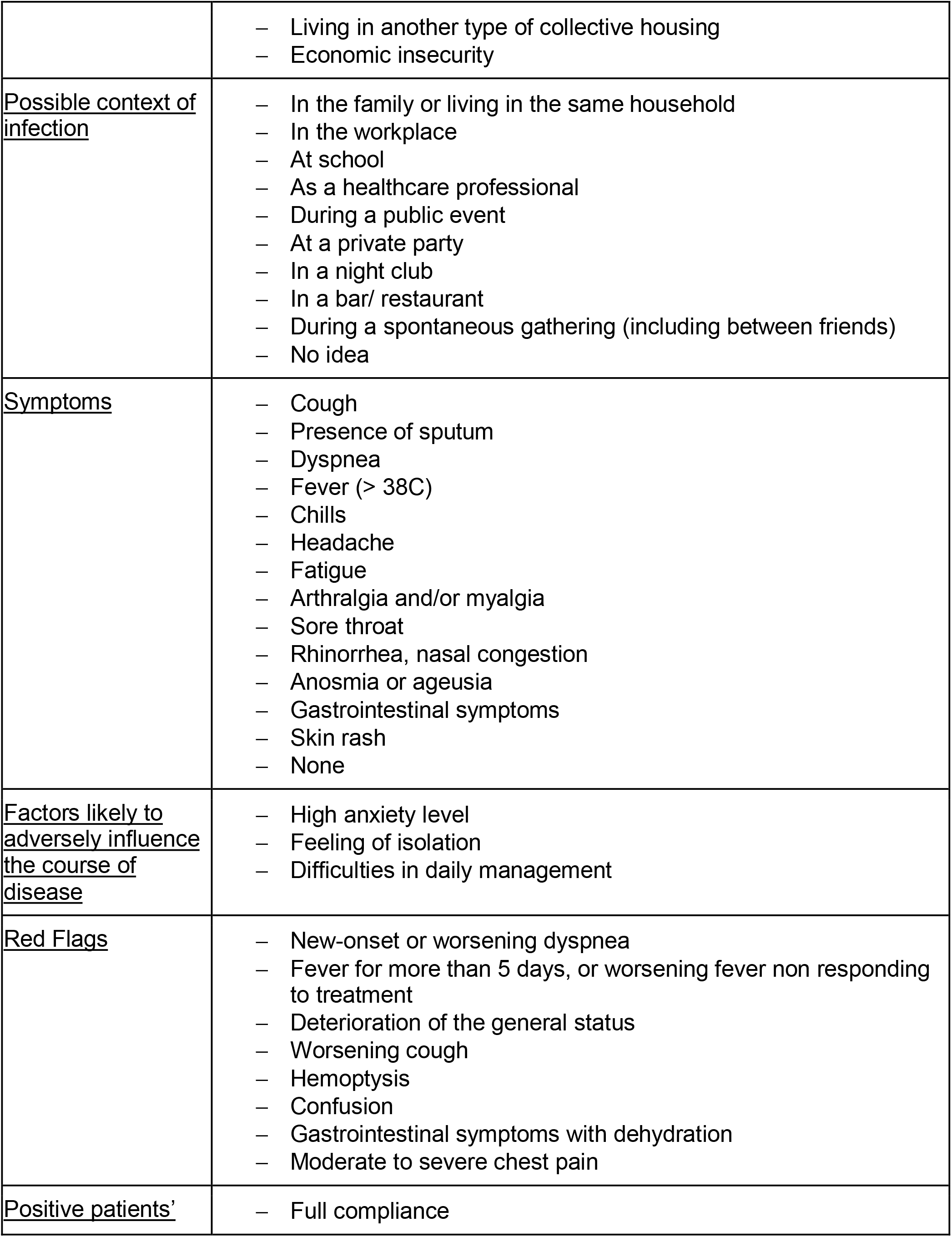
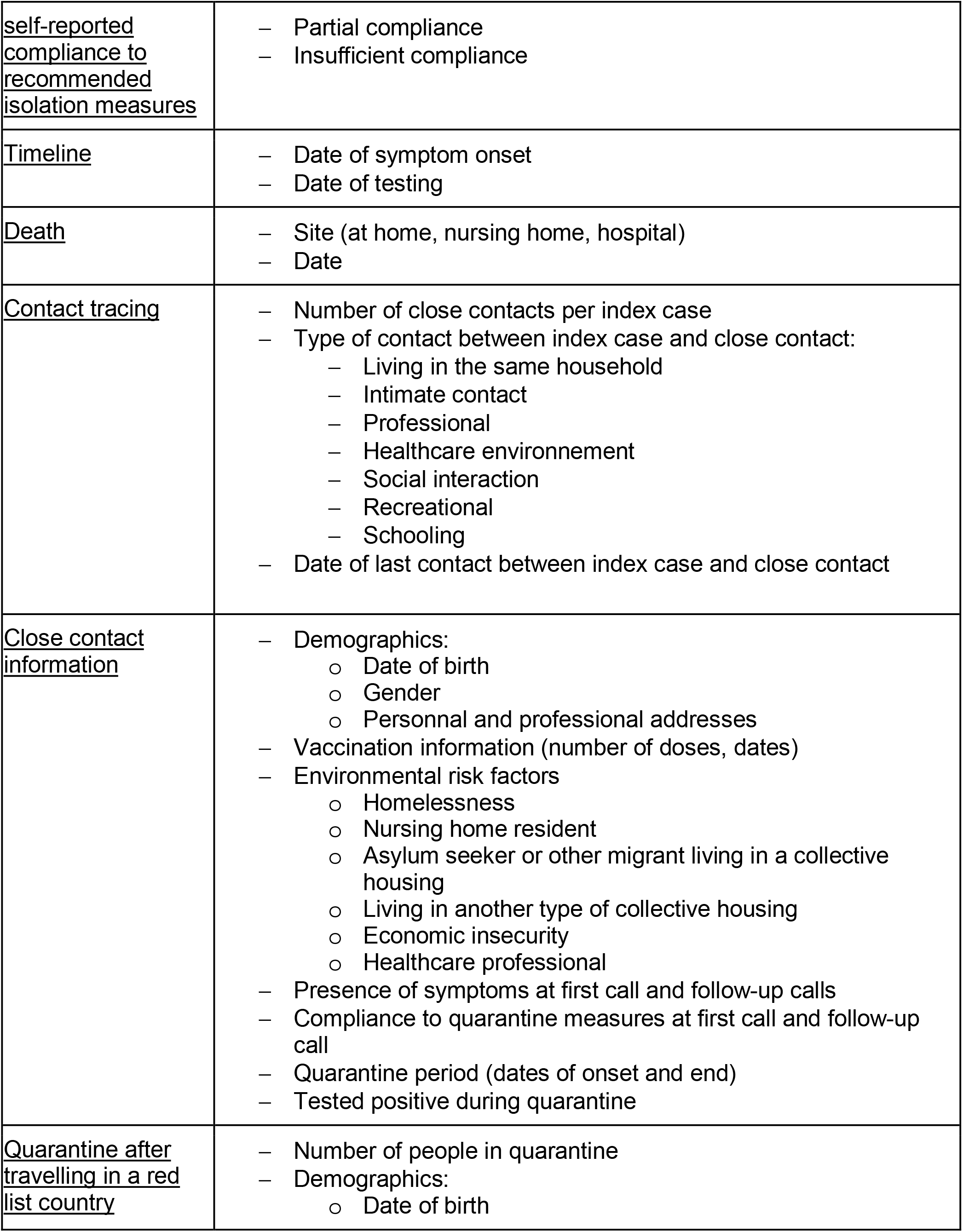
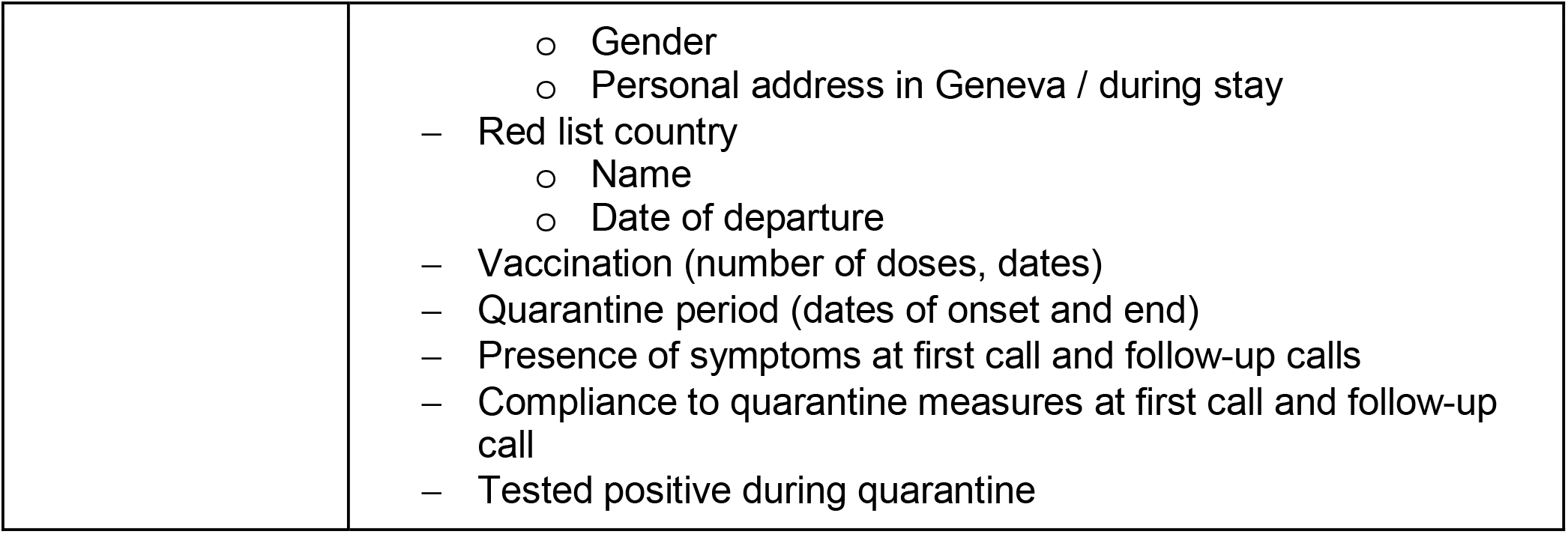
Actionable Register of Geneva Out- and inpatients with SARS-CoV-2 (ARGOS) collected data

## Findings to date

On June 1^st^, 2021, of all 360’525 patients recorded in the ARGOS database, 65’475 had at least one positive test result, 294’723 had one or more negative test results and no positive one, and 327 were suspected COVID-19 cases without a positive test to confirm the disease. During the same period, 655527 tests were performed, among which 89.2% were PCR. The positivity, i.e. the ratio between the positive tests and the total amount of tests, was of 10.7%. Among the positive patients, 4’687 persons did not allow their data to be used for research and were excluded from analyses. The remaining number of positive cases available for analysis is 60’788. Of these patients, 37.6% have only a first contact, 10.6% and 8.5% have one and two follow-up call respectively, and 27.7% of participants have three or more follow-up calls. 15.7% of the patients were not contacted, mainly during the periods of active pandemic activity when the GDH team was overworked (see Table 2). The cohort shows a slight female predominance, with women representing 50.2% to 55.9% of all patients depending on the defined period (Table 2). More than 60 percent of all recorded patients have no risk factor for a poor clinical outcome(18). The context of infection recorded for COVID19 positive patients since June 2020 indicates that infection mostly occurs at home, at work or via the educational system. Around 23.2% of the patient reported having no idea of their contamination context. Information about 114’690 close contacts of positive patients has been registered, and 639’153 days of quarantine have been notified. 9’551 close contacts of a positive COVID19 case had a positive test result during their quarantine. Given that the standard duration of a quarantine is 10 days, we can estimate that around 15% of the persons in quarantine after being in contact with a positive COVID19 case received a positive test result during their quarantine (see table 2).

**Table 2.** ARGOS baseline characteristics of positive patients, Geneva, February 26, 2020 – June 1st, 2021. Periods are presented by grouping together the first wave of cases, the period between the two waves, the second wave and the following period of sustained epidemic activity, and finally the more recent period following the start of the vaccination campaign.

|  | Overall | 2020-02-25<br>-> 2020-04-27 | 2020-04-27<br>-> 2020-09-24 | 2020-09-24<br>-> 2021-02-14 | 2021-02-14<br>->2021-06-02 |
| --- | --- | --- | --- | --- | --- |
| Number of positive patients |  |  |  |  |  |
| n | 60788 | 5782 | 3274 | 40882 | 10824 |
| Living in Geneva | 53344 (88.2) | 4793 (84.4) | 2827 (86.5) | 35936 (88.3) | 9775 (90.4) |
| Number of follow-up per patient recorded in ARGOS |  |  |  |  |  |
| Not called | 9514 (15.7) | 1135 (19.6) | 108 (3.3) | 8128 (19.9) | 131 (1.2) |
| First contact only | 22847 (37.6) | 3402 (58.8) | 578 (17.7) | 17541 (42.9) | 1316 (12.2) |
| 1 Follow-up call | 6427 (10.6) | 346 (6.0) | 735 (22.4) | 4387 (10.7) | 959 (8.9) |
| 2 follow-up calls | 5178 (8.5) | 152 (2.6) | 683 (20.9) | 2362 (5.8) | 1977 (18.3) |
| 3 or more follow-up calls | 16822 (27.7) | 747 (12.9) | 1170 (35.7) | 8464 (20.7) | 6441 (59.5) |
| Age |  |  |  |  |  |
| 0-19 | 6997 (11.5) | 175 (3.0) | 364 (11.1) | 4052 (9.9) | 2406 (22.2) |
| 20-39 | 21080 (34.7) | 1690 (29.2) | 1558 (47.7) | 14356 (35.1) | 3473 (32.1) |
| 40-64 | 23879 (39.3) | 2567 (44.4) | 1101 (33.7) | 16007 (39.2) | 4202 (38.8) |
| 65-80 | 5046 (8.3) | 676 (11.7) | 138 (4.2) | 3693 (9.0) | 539 (5.0) |
| >80 | 3750 (6.2) | 674 (11.7) | 108 (3.3) | 2769 (6.8) | 199 (1.8) |
| Gender |  |  |  |  |  |
| Male | 28314 (46.6) | 2549 (44.1) | 1628 (49.8) | 18890 (46.2) | 5238 (48.4) |
| Female | 32433 (53.4) | 3233 (55.9) | 1643 (50.2) | 21972 (53.8) | 5574 (51.5) |
| Non binary | 22 (0.0) | 0 (0.0) | 1 (0.0) | 8 (0.0) | 12 (0.1) |
| Comorbidities and risk factors |  |  |  |  |  |
| Cardiovascular disease | 1835 (3.0) | 396 (6.8) | 95 (2.9) | 1103 (2.7) | 241 (2.2) |
| Hypertension | 4469 (7.4) | 600 (10.4) | 196 (6.0) | 2968 (7.3) | 705 (6.5) |
| Diabetes | 1975 (3.2) | 273 (4.7) | 95 (2.9) | 1295 (3.2) | 312 (2.9) |
| Chronic respiratory illness | 2170 (3.6) | 512 (8.9) | 83 (2.5) | 1269 (3.1) | 306 (2.8) |
| kidney | 229 (0.4) | N/A | N/A | 186 (0.5) | 43 (0.4) |
| Cancer | 545 (0.9) | 73 (1.3) | 28 (0.9) | 349 (0.9) | 95 (0.9) |
| Immunosuppression | 600 (1.0) | 192 (3.3) | 30 (0.9) | 301 (0.7) | 77 (0.7) |
| obesity | 1081 (1.8) | N/A | N/A | 778 (1.9) | 303 (2.8) |
| Age 65 and older | 8796 (14.5) | 1350 (23.3) | 246 (7.5) | 6462 (15.8) | 738 (6.8) |
| No risk factor | 36905 (60.8) | 1868 (32.4) | 2523 (77.1) | 24093 (59.0) | 8419 (77.8) |
| Missing information | 8698 (14.3) | 1854 (32.1) | 209 (6.4) | 6221 (15.2) | 411 (3.8) |
| Other potential risks |  |  |  |  |  |
| Chronic disease | 787 (1.3) | 56 (1.0) | 28 (0.9) | 550 (1.3) | 152 (1.4) |
| smoking | 4659 (7.7) | N/A | N/A | 3345 (8.2) | 1313 (12.1) |
| pregnancy | 533 (0.9) | 41 (0.7) | 24 (0.7) | 352 (0.9) | 116 (1.1) |
| Other risk | 4284 (7.0) | 107 (1.9) | 364 (11.1) | 2741 (6.7) | 1072 (9.9) |
| Self reported symptoms |  |  |  |  |  |
| Missing information | 12735 (21.0) | 2632 (45.5) | 264 (8.1) | 9217 (23.3) | 604 (5.0) |
| no symptoms ever declared | 3893 (6.4) | 254 (4.4) | 416 (12.7) | 1807 (4.6) | 1413 (11.6) |
| At least one symptom | 44159 (72.6) | 2896 (50.1) | 2594 (79.2) | 28516 (72.1) | 10148 (83.4) |
| Possible context of infection |  |  |  |  |  |
| family | 17266 (28.4) | N/A | 511 (15.6) | 11861 (29.0) | 4889 (45.2) |
| work | 8535 (14.0) | N/A | 304 (9.3) | 6588 (16.1) | 1639 (15.1) |
| school | 3302 (5.4) | N/A | 0 (0.0) | 2200 (5.4) | 1101 (10.2) |
| Health care worker | 894 (1.5) | N/A | 17 (0.5) | 808 (2.0) | 67 (0.6) |
| Public event | 204 (0.3) | N/A | 22 (0.7) | 138 (0.3) | 44 (0.4) |
| private_party | 1372 (2.3) | N/A | 184 (5.6) | 933 (2.3) | 255 (2.4) |
| club | 70 (0.1) | N/A | 24 (0.7) | 41 (0.1) | 5 (0.0) |
| restaurant | 1346 (2.2) | N/A | 161 (4.9) | 1125 (2.8) | 60 (0.6) |
| Spontaneous gathering | 2527 (4.2) | N/A | 81 (2.5) | 1718 (4.2) | 728 (6.7) |
| No idea | 14090 (23.2) | N/A | 410 (12.5) | 10451 (25.6) | 3222 (29.8) |
| Other | 4921 (8.1) | N/A | 488 (14.9) | 3574 (8.7) | 858 (7.9) |
| Missing information | 16520 (27.2) | 5775 (100) | 1356 (41.4) | 8885 (21.7) | 486 (4.5) |
| Profession |  |  |  |  |  |
| health care professional | 4503 (7.4) | 902 (15.6) | 175 (5.3) | 2973 (7.3) | 452 (4.2) |
| Environmental risk factor |  |  |  |  |  |
| Homelessness | 135 (0.2) | 15 (0.3) | 6 (0.2) | 102 (0.2) | 12 (0.1) |
| Nursing home resident | 1895 (3.1) | 377 (6.5) | 63 (1.9) | 1403 (3.4) | 49 (0.5) |
| Asylum seeker or other migrant living in a collective home | 267 (0.4) | 25 (0.4) | 2 (0.1) | 172 (0.4) | 68 (0.6) |
| Collective home resident (other than migrant) | 627 (1.0) | 34 (0.6) | 31 (0.9) | 431 (1.1) | 131 (1.2) |
| Reason for testing |  |  |  |  |  |
| Acute symptoms | 45321 (88.0) | 5633 (99.9) | 2693 (85.9) | 28785 (89.2) | 8204 (78.8) |
| Testing |  |  |  |  |  |
| Total number of tests performed | 655527 | 28931 | 80342 | 291510 | 254744 |
| PCR | 584573 (89.2) | 28879 (99.8) | 80339 (100.0) | 263182 (90.3) | 212173 (83.3) |
| Number of patient tested | 360525 | 25853 | 71269 | 210598 | 169164 |
| Positivity rate | 9.4 | 21.0 | 4.1 | 14.3 | 4.3 |
| deaths |  |  |  |  |  |
| Deaths number | 747 | 280 | 20 | 421 | 22 |
| Age | 87.1 [80.2, 91.5] | 86.3 [79.4, 91.3] | 89.2 [85.8, 93.3] | 87.7 [81.4, 91.8] | 83.6 [70.5, 90.3] |
| Gender | 354 (47.4) | 130 (46.4) | 10 (50.0) | 200 (47.7) | 11 (45.8) |
| Contact tracing |  |  |  |  |  |
| Number total of contact | 114690 | 118 | 12420 | 77990 | 24162 |
| Number of contact per index patient | 3 [1, 6] | 0 [0, 0] | 7 [4, 11] | 3 [1, 6] | 3 [2, 5] |
| Quarantine after contact with positive |  |  |  |  |  |
| Number of days | 639153 | N/A | 31615 | 445468 | 162003 |
| Number of infection during quarantine | 9551 | N/A | 333 | 6009 | 3209 |
| Pourcentage of quarantine leading to infection | 14.94 | N/A | 10.53 | 13.49 | 19.81 |
| Quarantine after traveling |  |  |  |  |  |
| Number of days | 273189 | N/A | 85490 | 121202 | 66429 |
| Number of infection during quarantine | 96 | N/A | 29 | 42 | 25 |
| Pourcentage of quarantine leading to infection | 0.35 | N/A | 0.34 | 0.35 | 0.38 |

273’189 days of quarantine concerning 27920 persons were ordered for persons coming back from a country at risk. These country were in order of importance Spain (19.4%), France (14.8%), Kosovo (7.6%), United States (7.0%), United Kingdom (7.0%), Portugal (6.2%) and Brazil (4.2%). 96 persons received a positive test result during their quarantine, among which 26 came back from Kosovo, 11 from France and 10 from Spain, the total of these infection occurring in 0.35% of the quarantines.

To mitigate confounding effects and improve data analysis and interpretation, we present the data according to four periods (see Figure 1).

**Figure 1.**
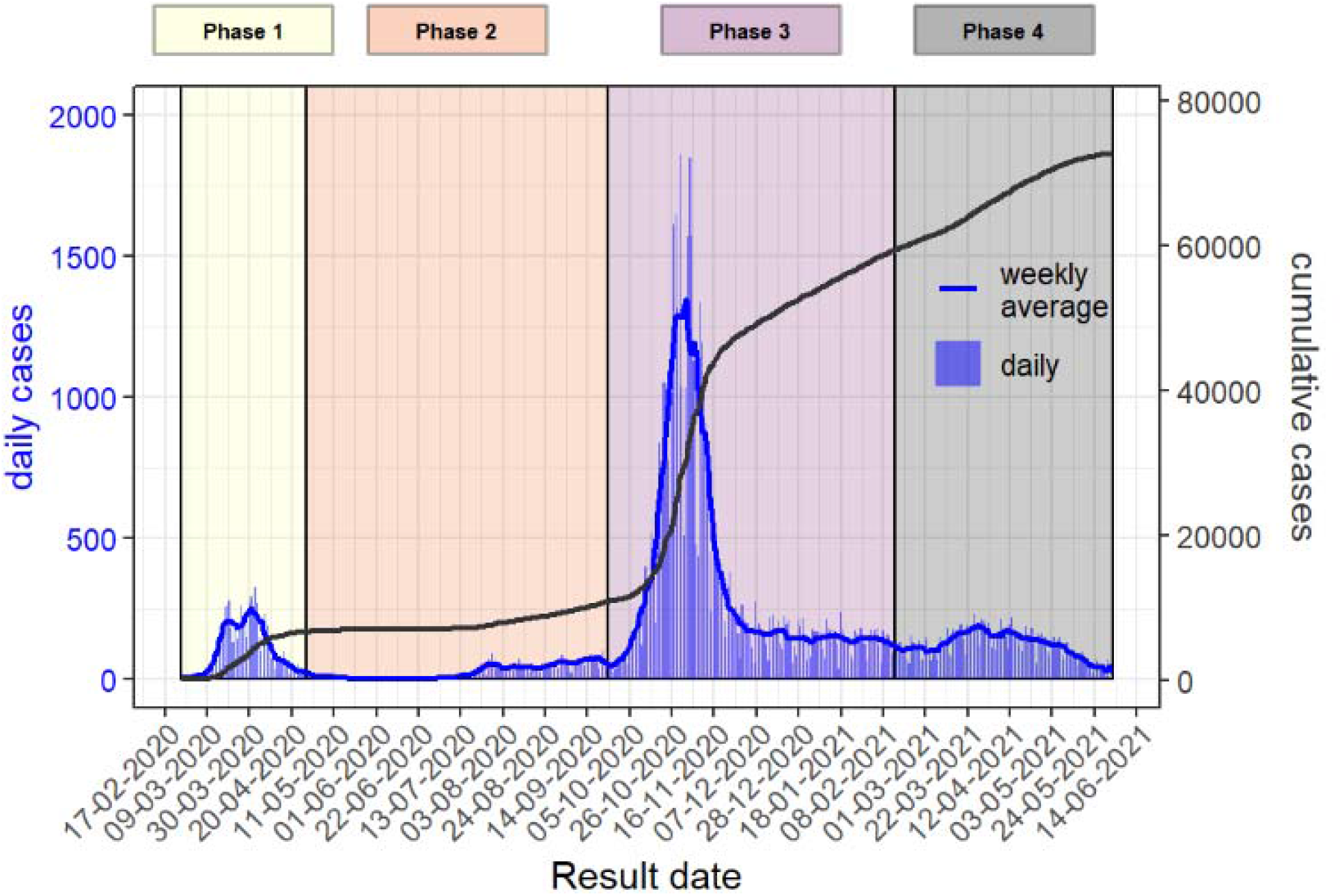
Epidemic Curve of the cases of Coronavirus Disease 2019 (COVID-19) in Geneva state, February 26, 2020 – June 1st, 2021. Vertical bars represent the daily cases (based on the date of the test result), solid blue line represents the weekly moving average and the solid black line the cumulative cases.

### February 26 to April 27, 2020 (first phase)

The first phase starts on February 26, 2020, when the first case was tested positive for SARS-CoV-2 in the Geneva area. The Swiss authorities implemented lockdown measures which remained moderate in comparison with many other countries (19). This first phase ends on April 27, 2020, when some of the measures started to be lifted following the decreasing incidence of new cases and hospitalizations. During this first wave, contact tracing was not implemented. Between March 13 and March 29, 2020, symptomatic individuals did not get tested due to shortage of testing materials, unless they were healthcare workers, considered at-risk or hospitalized. The percentage of healthcare professionals among positive cases was significantly higher during this phase (15.6%) and the percentage of patients declaring no risk factors was smaller (32.4%) when compared to the other phases. The positivity (i.e. the ratio between positive tests results and the total amount of test performed) was of 23%.

### April 28 to September 24, 2020 (second phase)

Between May and the end of September, 2020, incidence of SARS-CoV-2 positive cases remained low. Nearly all restrictions were lifted at the end of June. Nightclubs were closed again at the end of July after a surge of incidence mostly amongst Geneva youth, as can be seen by the relative higher incidence of the 20-39 year age category compared to the others during this period (Figure 2). 14.1% of the positive tests during this period were stemming from screening campaigns and more than 70% of cases reported no risk factor. The positivity was only 4.6% during this period.

**Figure 2.**
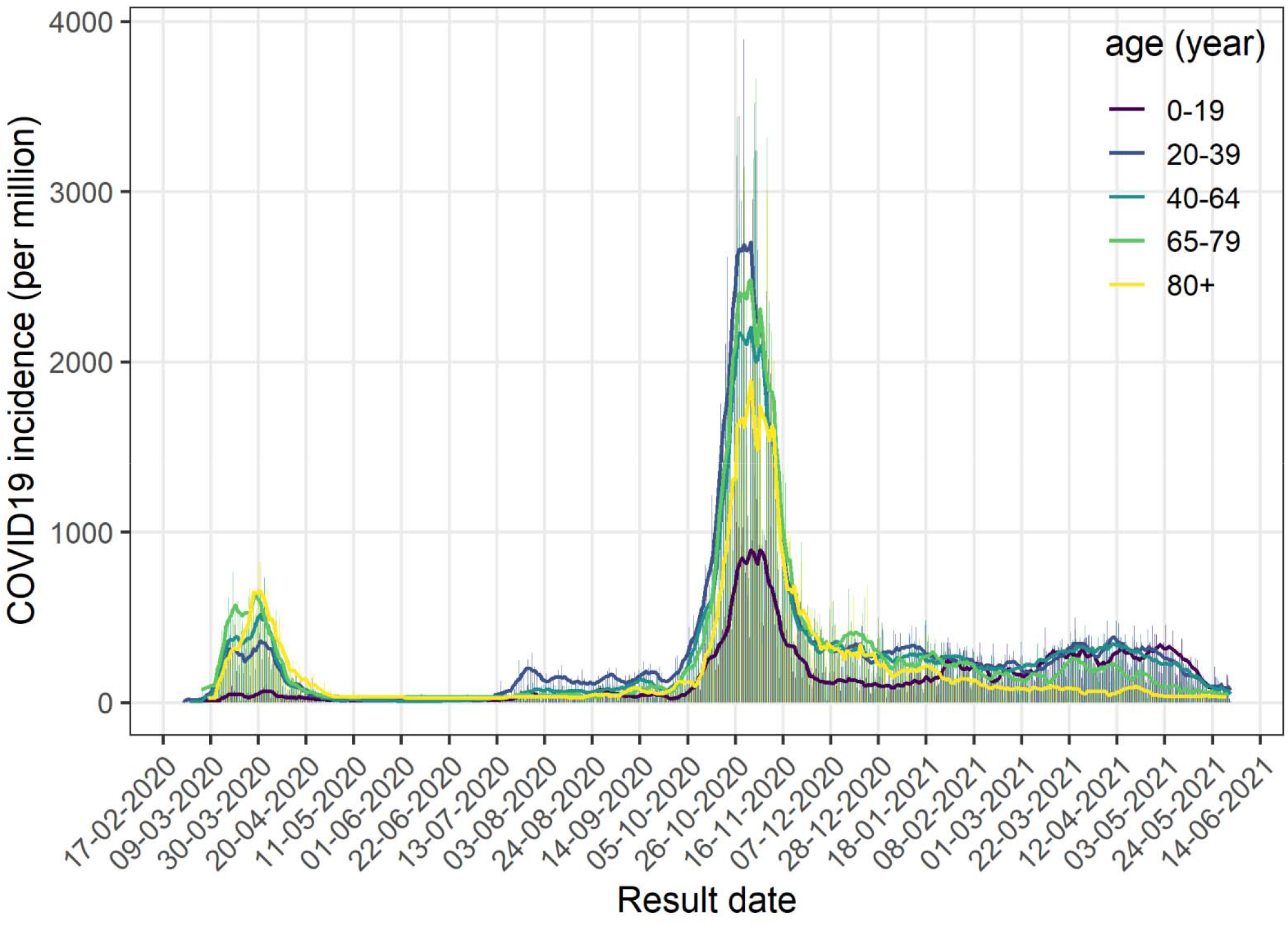
Incidence per age category, Geneva, February 26, 2020 – June 1st, 2021. Vertical bars represent the daily incidence, solid line represent the weekly moving average.

### September 24, 2020 to February 28, 2021 (third phase)

A second wave of SARS-CoV-2 positive cases hit Geneva in late September, 2020, at the same time as in the neighboring countries. New restrictions were imposed mid-October but no real lockdown was enacted. The peak lasted about 8 weeks. Due to political and economic pressure, some restrictions measures were lifted long before incidence reached low level. The number of SARS-CoV-2 positive cases in Geneva area remained significant during several months. During February 2021, the B.1.1.7 variant completely replaced SARS-CoV-2 wild type. At the same time, federal and local policies evolved and testing among symptomatic children over 5 years old was newly encouraged. Concurrently with these changes, the incidence of the 0-19 year age population almost doubled to reach those of the older age categories (19–21). The positivity during this period was 15.9%.

As of February 8^th^, 2021, quarantine measures for close contacts were lifted after 7 days if the person tested negative for SARS-CoV-2. Concurrently, the percentage of close contacts tested positive increased (see table 2). Considering vaccination program, the first dose of vaccine in Geneva was given to an elderly patient the 28^th^ December 2020. At first, only residents over 74 years old and patients with risk factors received vaccination. The decline of the incidence for people of the corresponding age category compared to the others can be observed since February 2021 in figure 2.

### March 1^st^ to 1^st^ June, 2021 (fourth phase)

On March 1^st^, access to vaccination continued to broaden. Resident over 65 years were allowed to be vaccinated since March 17, 2021, followed by the 45-65 year old residents starting at April 12, 2021. By 19 May, 2021, all Geneva residents over 15 years old were eligible to get vaccinated. The incidence COVID among the 65-79 year old population started to decline by end of March (figure 2), followed by a rapid decline of the incidence overall by mid-May. The amount of screening tests increased, as 21% of the positive tests were performed during a screening campaign. The positivity decreased to 5.2% during this period.

### Discussion

COVID-19 represents a major challenge to each country’s healthcare system. Collaboration between healthcare providers and public health authorities is particularly important in order to improve both our understanding of the disease and our response(22–24). The publication of the ARGOS cohort underscores our willingness to share data for research purposes. Indeed, data from this registry has already been used to investigate symptoms and long COVID (25), infection fatality rate (9), and re-infection rates (26), as well as viral load kinetics (27). Several projects using these data to develop more accurate mathematical models to estimate transmission chains are also ongoing.

Furthermore, analysis from the ARGOS database illustrates the impact of various testing policies on the proportion of risk factors or age groups identified among confirmed cases. The partition of data analysis and interpretation according to policy period confirms the variations within each group depending on the period of interest and could thus guide public health decisions.

## Strength and limitations

The state of Geneva accounts for half a million residents and the local Directorate of Health ordered the recording of all COVID-19 positive cases since the beginning of the epidemic, according to recommendations from the Federal Office of Public Health. Due to this policy, the database’s main strength consists of its large number of cases, representative of all diagnosed cases on a regional level primarily serving operational needs and not scientific purposes, with one main objective: assessing all cases. This cohort is also multicentric as it includes all tests performed in Geneva’s hospitals (both public and private), private practices and medical centers. The fact that a very large proportion of all cases are assessed reduces the risk of biased data. Also, as data is recorded on the day of the call to the patient, recall bias is very low. Finally, the ARGOS^3^ database is characterized by a high number of follow-ups.

Despite these strengths, ARGOS has been influenced by the testing policy and the results must be seen in light of these influences. First, individuals without risk factors for COVID-19 and those younger than 65 years old are underrepresented in the database during the testing restriction period. The shapes of the graphics in Figure 1 and 2 confirm the impact of this policy as there is a sudden decrease in number of cases after March 20, 2020, when restriction started. Other factors could have amplified this phenomenon such as less symptomatic forms of disease in younger people and children. Reasons to get tested have also evolved over the first months of the epidemic. For example, anosmia or ageusia became a testing criteria only in late April. Patients who presented with these isolated symptoms within the first two months of the epidemic could thus have been undertested. Seroprevalence study results confirm the underrepresentation of certain groups and the undertesting of the overall population (8).

Nevertheless, ARGOS has several limitations. First, measurement error due to lack of detail of some variables can be observed, since efficiency was prioritized over detail-oriented data collection. For instance, individuals’ level of education is not recorded. Secondly, misclassification also certainly occurs as symptoms and risk factors are self-reported. Moreover, recording of information in ARGOS is performed by a large and evolving team of professionals, including healthcare workers with various backgrounds, medical students, police recruits, or contact tracers with no particular medical and health knowledge. Due to the crisis situation, training contents delivered to the GDH team often evolved, leading to a certain level of heterogeneity of phone interviews and a greater risk for misclassification of medical information. Thirdly, the patient information gathered is tailored to operational needs and growing scientific knowledge. For example, anosmia and ageusia were initially classified as general ENT symptoms, and were later detailed separately as they were recognized as frequent and specific manifestations of COVID-19 (28). Finally, during some periods of the pandemic, the GDH team was overworked and could not call not verify self-reported information for all positive cases. This resulted in missing and incomplete data.

In conclusion, ARGOS is a large, real-world registry of individuals tested for SARS-Cov2^1^. Unlike many other registries, it involves every tested individual and is not limited to hospitalized patients, thus providing a precious resource to assess the impact of public health policies and overall disease burden of COVID-19.

## Data Availability

Deidentified ARGOS data can be available upon reasonable request, including a research protocol, using the form https://edc.hcuge.ch/surveys/?s=TLT9EHE93C.

## Collaboration

The publication of the ARGOS cohort underscores our willingness to share data for research purposes and for optimizing public health measures. Deidentified ARGOS data are available upon reasonable request, including a research protocol, using the following <u>form:</u> https://edc.hcuge.ch/surveys/?s=TLT9EHE93C.

## Further details

### Data sharing statement

The deidentified data underlying this article will be shared on reasonable request using the <u>form</u> (https://edc.hcuge.ch/surveys/?s=TLT9EHE93C)

### Ethics approval

Research received the agreement of the Cantonal Ethic Committee of Geneva (CCER protocol 2020-01273).

### Funding

ARGOS is supported by Geneva State public funds and the research project SELFISH financed by the Swiss National Science Foundation LIVES, grant number 51NF40-160590.

## Acknowledgements

We thank all members of the COVID-19 team at the General Directorate of Health, as well as all healthcare providers and laboratories involved in the management of COVID-19 patients. We also wish to thank all patients and their contacts who are included in the ARGOS database.

## Authors contribution

Each author contributed to this article, based on the criteria of the International Committee for Medical Journal Editors. Camille Genecand conceptualized, designed the article format, interpreted the data, and conducted the literature review. Denis Mongin conducted the data analysis and participated in its formulation and its interpretation in the text. Flora Koegler conceptualized and designed the article format, interpreted the data, and conducted the literature review. Dan Lebowitz participated to the article design and reviewed it. Delphine Courvoisier designed the study’s analytic strategy, reviewed the article, and revisited it critically. Simon Regard, Pierre Chopard, Marwène Grira, Elisabeth Delaporte, Mayssam Nehme, Olivia Braillard, Dominique Joubert, Idris Guessous, Jerome Stirnemann and Aglaé Tardin helped acquisition of data and reviewed the article’s content critically.

## Footnotes

1 Severe acute respiratory syndrome coronavirus 2

2 coronavirus disease 2019

